# Limited Utility of Cardiovascular Risk Scores for People Living with HIV in Malawi

**DOI:** 10.1101/2020.08.01.20166462

**Authors:** Chia Y Goh, Henry Mwandumba, Alicja Rapala, Willard Tinago, Irene Sheha, Mishek Chammudzi, Patrick Mallon, Nigel Klein, Saye Khoo, C Kelly

**Author notes:** <u>Correspondence</u> Christine Kelly. <u>Conference presentations</u> Infectious Diseases Society of Ireland Annual Scientific Meeting, Galway, May 2017.

## Abstract

HIV is associated with increased cardiovascular disease (CVD) risk. Despite the high prevalence of HIV in low income sub-Saharan Africa, there are few data on the assessment of CVD risk in the region. In this study, we aimed to compare the utility of existing CVD risk scores in a cohort of Malawian adults, and assess to what extent they correlate with established markers of endothelial damage: carotid intima-media thickness (IMT) and pulse wave velocity (PWV).

WHO/ISH, SCORE, FRS, ASCVD, QRISK2 and D:A:D scores were calculated for 279 Malawian adults presenting with HIV and low CD4. Correlation of the calculated 10-year CVD risk score with IMT and PWV was assessed using Spearman’s rho.

The median (IQR) age of patients was 37 (31 – 43) years and 122 (44%) were female. Median (IQR) blood pressure was 120/73mmHg (108/68 – 128/80) and 88 (32%) study participants had a new diagnosis of hypertension. The FRS and QRISK2 scores included the largest number of participants in this cohort (96% and 100% respectively). D:A:D, a risk score specific for people living with HIV, identified more patients in moderate and high-risk groups. Although all scores correlated well with physiological markers of endothelial damage, FRS and QRISK2 correlated most closely with both IMT [r^2^ 0.51, p<0.0001 and r^2^ 0.47, p<0.0001 respectively] and PWV [r^2^ 0.47, p<0.0001 and r^2^ 0.5, p<0.0001 respectively].

Larger cohort studies are required to adapt and validate risk prediction scores in this region, so that limited healthcare resources can be effectively targeted.

## Introduction

Non-communicable diseases (NCDs) are responsible for 85% of premature deaths in low and middle income countries, and are predicted to overtake those from infectious comorbidities by 2030 (1, 2). In sub-Saharan Africa, NCDs are projected to cost low and middle-income countries (LMICs) 17 trillion dollars over the next 15 years (3). At the same time, the number of people living with HIV (PLWH) aged over 50 years in LMICs has nearly tripled over the past 20 years, currently including over three million people in Eastern and Southern Africa (4). Cardiovascular events such as myocardial infarction and strokes are increased by approximately two fold amongst people living with HIV in high income settings (5). In Malawi, people presenting with low CD4 counts are over 15 times more likely to have a stroke (6). The relationship between markers of HIV disease and inflammation driven cardiovascular disease is well established; unsuppressed HIV viral loads, low nadir CD4 counts and markers of inflammation such as IL-6, Ddimer and CRP are all associated with higher risk of cardiovascular events amongst people living with HIV (7-9).

In a context where healthcare systems are structured around management of acute infectious episodes, integration of chronic illness care is challenging. HIV management infrastructure poses a promising framework upon which to build chronic disease management systems in sub-Saharan Africa (10). However, the paucity of data on NCDs in the region causes difficulty in policy making and priority setting (11). It also makes risk stratification difficult in the clinical setting, where established tools are needed to identify those patients with higher cardiovascular risk, so that limited resources in the region can be efficiently targeted.

Currently available cardiovascular risk scores have been shown to underestimate cardiovascular risk amongst HIV populations in high income countries (12). Recommendations on which scores to use amongst HIV populations in high income settings vary. The British HIV Association recommends QRISK (13), The European AIDS Clinical Society recommends the Framingham Risk score (14), and the Infectious Diseases Society of America references the Atherosclerotic Cardiovascular Disease (ASCVD) score (15). These scores have been validated in European and American populations respectively which likely influences the choice of score. D:A:D is the only score developed in a HIV population and accounts for HIV-related factors (16), but has been less robustly validated. SCORE and WHO tools have been adapted to be more practical in a low resource setting, but these are only validated amongst high income cohorts (17, 18). The scarcity of data from sub-Saharan Africa means that there are no cardiovascular risk scores currently validated for these populations (19).

With this study, we applied these risk scores to a sub-Saharan Africa population with advanced immune suppression (and therefore higher long term risk of cardiovascular events), aiming to assess their utility and compare them with physiological markers of endothelial damage.

## Methods

### Study design and Population

We conducted a cross-sectional study of adults living with HIV and aged over 18 years recruited from Queen Elizabeth Central Hospital, Blantyre. Participants had a CD4 count less than 100 cells/uL and were enrolled 2 weeks following antiretroviral therapy (ART) initiation. A detailed clinical evaluation was carried out, including assessment of cardiovascular risk factors, blood draw for total cholesterol and fasting blood sugar, and non-invasive measurements of endothelial dysfunction.

All participants provided informed written consent and ethical approval was granted by the College of Medicine Research and Ethics Committee (COMREC), University of Malawi (P.09/13/1464) and the University of Liverpool Research and Ethics Committee (UoL000996).

### Cardiovascular risk scores

Six cardiovascular risk scores were assessed:(i) Atherosclerotic Cardiovascular Disease score (ASCVD), (ii) Framingham Risk Score (FRS), (iii) QRISK2, (iv) D:A:D score, (v) Systematic Coronary Risk Evaluation (SCORE) tool, and (vi) World Health Organisation/ International Society of Hypertension (WHO/ISH) score. A summary of the components of each of these scores is presented in Figure 1A.

**Figure 1.**
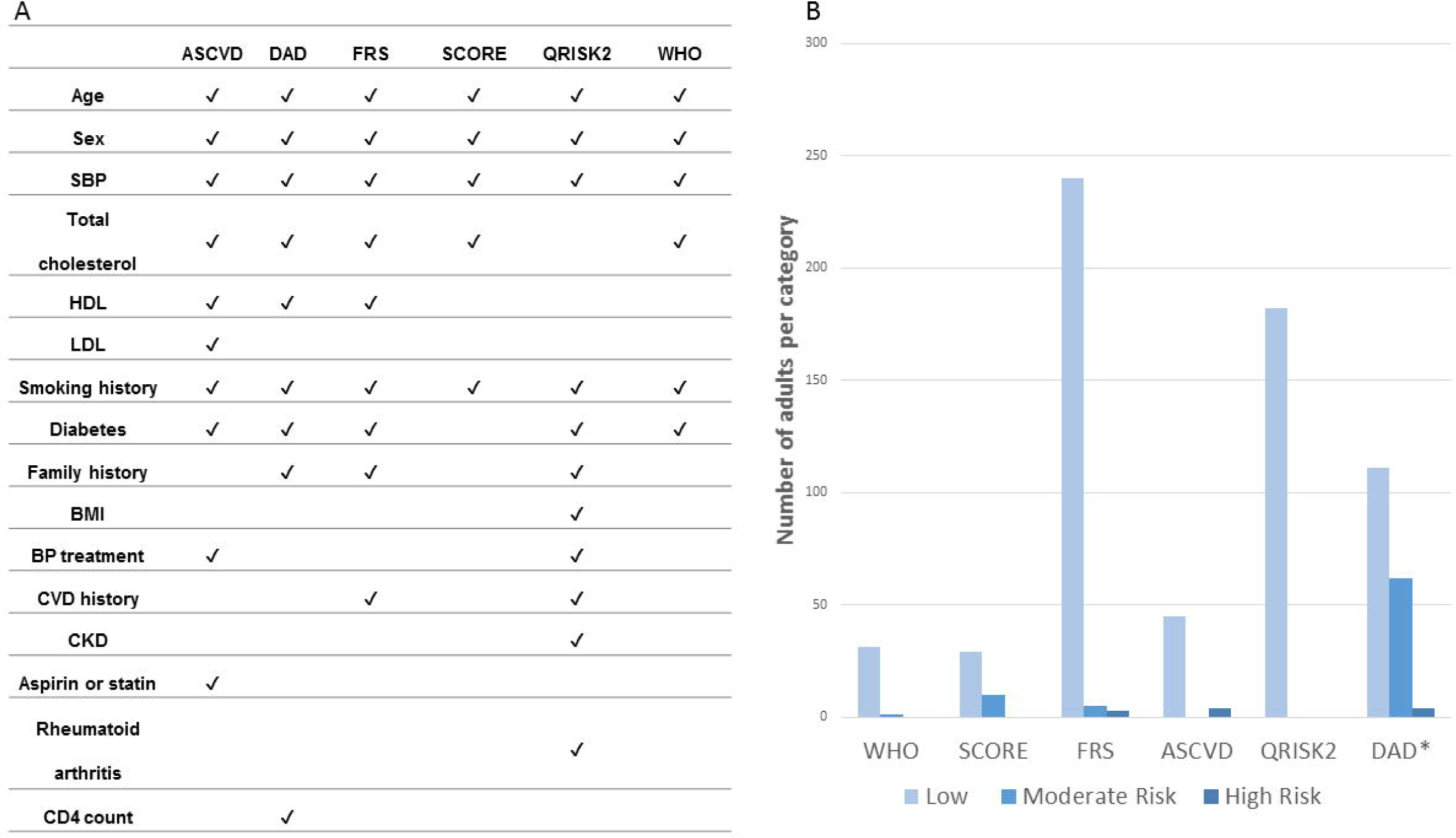
Score components and categorization of cardiovascular risk for 279 Malawian Adults living with HIV. **A**. Summary of variables included in each cardiovascular risk score. **B**. The number of participants classified as low, moderate and high risk of cardiovascular events. *All scores report a 10-year cardiovascular disease risk, apart from D:A:D which reports a 5-year risk.

Scores were individually calculated using the clinical data collected and presented as a percentage risk of experiencing a cardiovascular event in the next 10 years. The D:A:D score is the only exception and gives the event risk for a 5-year period.

We did not have HDL measurements available. Due to the normal total cholesterol values in this cohort, and the low incidence of dyslipidaemia reported in low income sub-Saharan Africa populations, we entered HDL values in the mid-range of normal to allow risk calculation for those scores that required it. However, for other missing variables, it was necessary to exclude the participants who did not meet score criteria. For the diagnosis of new hypertension, a cut off above 140/90 was used in accordance with American Heart Association and European Society of Cardiology guidelines (20, 21).

### Markers of endothelial damage

Carotid femoral Pulse Wave Velocity (PWV) was used to assess large vessel arterial stiffness and was measured using a Vicorder device (Skidmore Medical, London, UK). The distance was taken as the length from the sternal notch to the umbilicus and then the top mid-point of the femoral cuff, multiplied by 0.8 as per consensus guidelines (22, 23). Wave forms were saved, and a random sample was reviewed at three timepoints during the study to ensure consistent quality.

Carotid intima-media thickness (IMT) was measured subclinical atherosclerosis using B mode ultrasound (SIUI CTS 7700, Trisonics) with a 7hz linear array transducer. Three images were captured at the common carotid artery bilaterally, at least 10mm from the bifurcation. The intima media thickness at each site was determined using semi-automated edge detection software (Carotid Analysis for Research software, Mia-IIc, Iowa, USA) following a standardised protocol (24). A random sample of images were re-analysed by a second operator, who was blind to HIV status.

### Statistical analysis

The number of participants scored as moderate or high risk was compared using the chi square test. Correlations between 10-year cardiovascular event risk and physiological markers of endothelial damage were measured using Spearman rank sum. Analysis was undertaken using Stata 13.1 (Statacorp, College Station, USA).

## Results

### Clinical characteristics

The cohort consisted of 279 participants living with HIV with median (IQR) age 37 (31 – 43) and 122 (44%) female. 136 (53%) had attained a maximum of primary school education. Median (IQR) values for traditional risk factors were 120/73 mmHg (108/68 – 128/80) for blood pressure; 20 kg/m^2^ (18 – 22) for BMI; 3.6 mmol/L (3.0 – 4.4) for fasting total cholesterol; 4.9 mmol/L (4.4 – 5.6) for fasting glucose; and 65 µmol/L (54 – 78) for creatinine. The number of people who smoked cigarettes currently or in the past was 56 (20%). One participant had an existing diagnosis of diabetes at study enrolment. Five (2%) had a diagnosis of hypertension prior to study enrolment and were taking anti-hypertensives. Notably, a remarkable 88 (32%) study participants met the criteria for a new diagnosis of hypertension during the study period. The median (IQR) age for those with a new diagnosis of hypertension was 40 (34 – 47) years. No participants were on aspirin or statin, had rheumatoid arthritis, or had a family history of cardiovascular disease. The median (IQR) CD4 count was 41 (18 - 62) cells/µL and HIV viral load was 5.06 (4.62 – 5.47) (log10 copies/mL).

### 5- or 10-year cardiovascular event risk

SCORE, ASCVD, FRS, WHO/ISH and QRISK2 reported a 10-year cardiovascular event risk, whereas D:A:D reported a 5-year risk. Overall, the proportion of patients classified into moderate or high risk categories varied widely. Amongst those for whom the scores could be applied, the proportion of patients with a moderate or high risk of a cardiovascular event was: D:A:D 140(37%), SCORE 12(24%), ASCVD 3(8%), FRS 14(4%), WHO/ISH 1(2%), QRISK2 0(0%) (Figure 1B). The number of patients classified as high risk by the D:A:D score – which incorporates CD4 count - was significantly higher than ASCVD (p=0.006), FRS (p=0.04), QRISK2 (p<0.0001) and WHO (<0.0001), but not SCORE (p=0.20).

### Exclusion from scoring

Of the six scores assessed, five had criteria where the upper or lower bounds of at least one parameter rendered the score not applicable to study participants. The SCORE tool excluded the most participants, with 330 (85%) not eligible for assessment (246 [63%] younger than 40 years, 1 [0.3%] older than 69 years, 45 [12%] with systolic blood pressure less than 120mmHg, and 38 [10%] with total cholesterol less than 4.0mmol/L). The WHO/ISH score excluded 329 (85%) of participants for similar reasons (246 [63%] younger than 40 years, 45 [12%] with systolic blood pressure less than 120mmHg, and 38 [10%] with total cholesterol less than 4.0mmol/L). For the ASCVD score, 246 (71%) of cohort participants could not be assessed (256 [63%] younger than 40 years and 29 [8%] total cholesterol less than 3.2mmol/L). The FRS excluded 38 (10%) of patients from assessment due to total cholesterol less than 4.0mmol/L, and QRISK2 excluded 27 (7%) patients due to age younger than 25 years. Notably, all patients were eligible for assessment with the D:A:D score.

### Correlation of cardiovascular disease risk scores with endothelial damage

At enrolment, the median(IQR) carotid femoral PWV of the patients was 7.3 (6.5-8.2) m/s, and the median(IQR) common carotid IMT was 0.59 (0.56-0.67) mm. All scores correlated significantly with both PWV and IMT (see figure 2). FRS and QRISK2 correlated most closely over both modalities (PWV: r^2^ 0.47, p<0.0001 and r^2^ 0.5, p<0.0001 respectively; IMT: r^2^ 0.51, p<0.0001 and r^2^ 0.47, p<0.0001 respectively). Comparisons with the D:A:D score were limited by the fact that it reports a 5-year risk and that a small number of outliers demonstrated extreme high scores.

**Figure 2.**
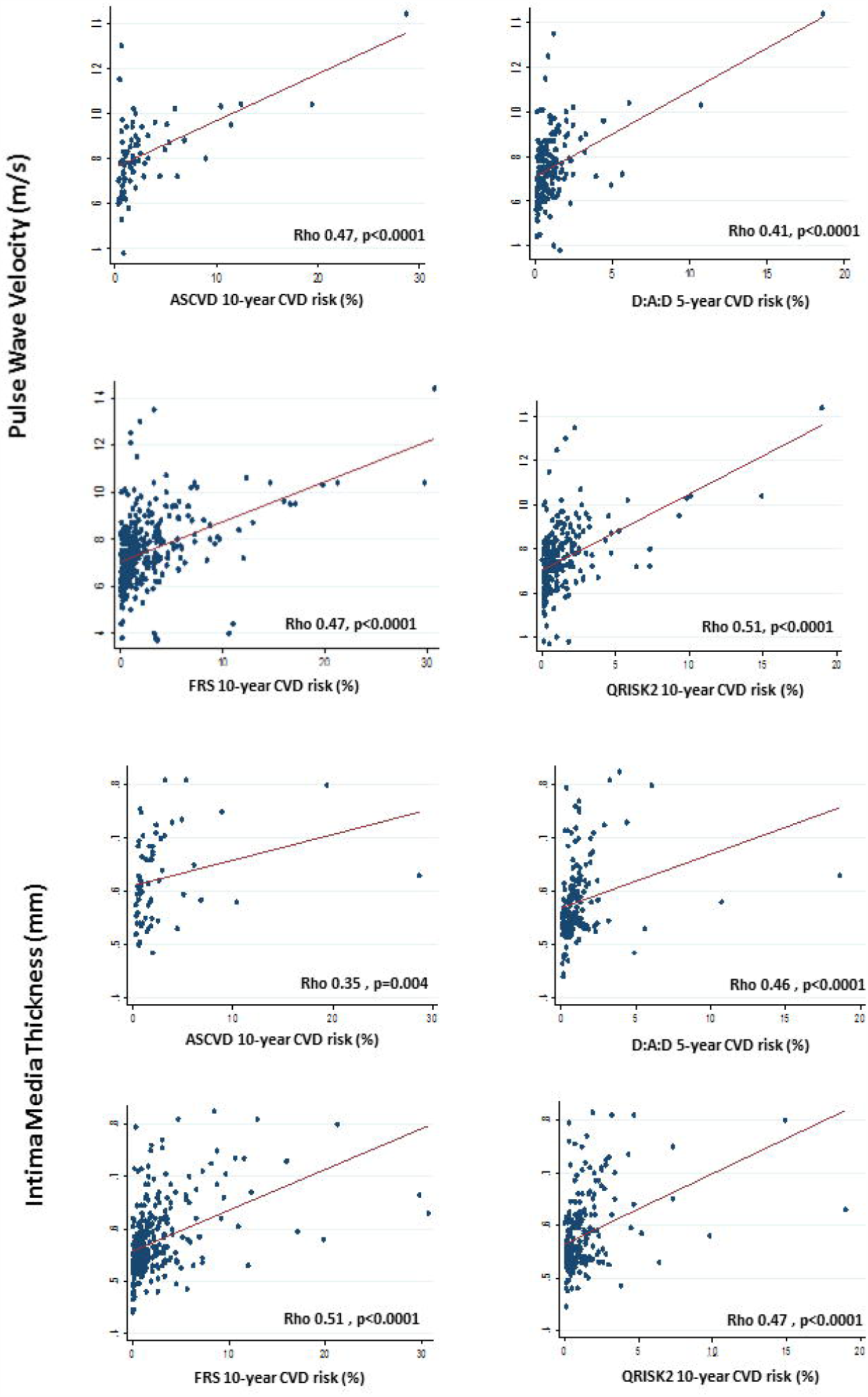
Correlation of CVD risk score with Pulse Wave Velocity and Intima-Media Thickness. 10-year cardiovascular risk (or 5-year for D:A:D) is plotted against Pulse Wave Velocity (PWV) and Intima-Media Thickness (IMT) values. The correlation of CVD risk score with PWV and IMT are presented using spearman rho. Correlations were not tested for SCORE and WHO/ISH due to the low number of participants assessed by these scores.

## Discussion

Many of the most commonly used cardiovascular risk scores demonstrate limited applicability in this cohort of Malawian adults presenting with HIV. For participants who could be assessed, estimates of cardiovascular risk varied, but correlated similarly with physiological markers of endothelial damage.

Colleagues in Tanzania showed higher cardiovascular risk amongst people living with HIV as assessed by the ASCVD score (25). Mosepele *et al* compared the ASCVD to FRS amongst a population in South Africa, and demonstrated that the ASCVD classified more patients with a high cardiovascular disease risk, but highlighted that a region specific score is required (26). Interestingly, a Ugandan study examining cardiovascular risk showed a pragmatic modified version of the FRS correlated closely with other risk scores (29).

As demonstrated here, cohorts of patients living with HIV in low income sub-Saharan Africa are relatively young, but rates of hypertension are remarkably high. But for some scores, an exclusion was applied to those aged less than 40 years. It is well documented that patients living with HIV experience chronic comorbidities, traditionally associated with older cohorts, at younger ages (30). This is likely to be no less true in low income sub-Saharan Africa, where risk factors for infection driven chronic inflammation are prevalent, and where urbanisation is leading to increases in other traditional risk factors that have, until recently, been uncommon amongst these populations (31).

Even in high income settings, currently available risk scores are not taking HIV related risk factors into consideration. The D:A:D score includes an assessment of CD4 count and anti-retroviral therapy regime but requires updates to include more recent modern regimes and markers of inflammation. The QRISK3 score, which has recently been launched, builds on previous versions to incorporate more risk factors for chronic inflammation. This exemplifies the importance of considering the inflammatory component of HIV when developing these risk predictions, and a similar process could be followed to render scores applicable to patients in low income sub-Saharan Africa.

Estimates of cardiovascular risk varied across scores, but correlated similarly with both arterial stiffness and subclinical atherosclerosis. FRS and QRISK2 correlated well with both arterial stiffness and subclinical atherosclerosis over a range of values and included a large proportion of participants. These may be the best available scores for use amongst HIV cohorts in this setting whilst awaiting the development of a better alternative.

This study was limited by the lack of a clinical outcome measure to validate the scores. We assessed a relatively small number of people with advanced immune suppression and correlations with arterial stiffness and subclinical atherosclerosis may be skewed by outliers with higher cardiovascular risk. However, we assessed a broad range of cardiovascular scores amongst a carefully characterized cohort of patients and linked these with validated physiological markers of endothelial damage.

Large longitudinal cohorts examining cardiovascular disease events are urgently required in low income sub-Saharan Africa. The design and validation of appropriate risk prediction scores relevant to people living with HIV will help target limited resources towards reducing morbidity and mortality from cardiovascular diseases.

## Data Availability

All relevant data is published with the manuscript

## Acknowledgement

The authors would like to acknowledge the patients and their families, as well as the staff in the ART clinic and Department of Medicine at Queen Elizabeth Central Hospital, Blantyre, Malawi.

We would like to thank the REALITY trial group for their support with study design and implementation.

